# Impact of immunization programs on 11 vaccine-preventable diseases in China: 1950-2018

**DOI:** 10.1101/2020.07.01.20144246

**Authors:** Jinhua Pan, Yesheng Wang, Zundong Yin, Zhixi Liu, Ying Wang, Qi Zhao, Weibing Wang

**Affiliations:** Fudan University, School of Public Health, Department of Epidemiology, Shanghai, China; School of Public Health, Key Laboratory of Public Health Safety of Ministry of Education, Fudan University, Shanghai 200032, China; National Immunization Program, Chinese Center for Disease Control and Prevention, Beijing 100050, China; Fudan University, School of Public Health, Department of Social Medicine, Shanghai, China

**Author notes:** These authors contributed equally to this work (Jinhua Pan, Yesheng Wang). Corresponding author: Weibing Wang. This study was funded by the “Evaluation project on national immunization program of China”.

**Keywords:** China, Vaccine-preventable diseases, Morbidity, Mortality, Immunization

## Abstract

**Introduction:** China’s immunization program has caused dramatic declines in morbidity and mortality from vaccine-preventable diseases (VPDs). To further evaluate the program’s achievements, we explored patterns in the morbidity and mortality of VPDs in China based on national surveillance data.

**Methods:** According to the times at which vaccines were introduced and the history of the National Immunization Program (NIP), we divided the study period of 1950 to 2018 into four stages. Meningococcal meningitis, measles, polio, diphtheria, pertussis, tuberculosis, hepatitis B, hepatitis A, Japanese encephalitis (JE), mumps and rubella were analyzed as representative notifiable diseases in China, and annual surveillance data obtained between 1950 and 2018 were used to derive trends in the morbidity and mortality rates of the 11 diseases for each stage. Quasi-Poisson regression models were used to estimate the impacts of specific vaccine programs, and life-table methods were used to calculate the loss of life expectancy, years of life lost (YLLs) and loss of working years.

**Results:** The total notification number for the 11 VPDs was 211,866,000 from 1950 to 2018. The greatest number occurred in 1959, with around 12 million cases and a total incidence rate of 1723 per 100,000 persons. The quasi-Poisson regression models showed that most of the 11 diseases revealed dramatic declines in both morbidity and mortality after a vaccine became available. From 1978 to 2018, the incidence of pertussis fell 98% from 126.35 to 1.58 per million, that of measles fell 99% from 249.76 to 0.28 per 100,000, that of meningococcal meningitis fell 99% from 32.23 to 0.01 per million, that of JE fell 98% from 5.39 to 0.13 per million, and diphtheria and polio were eradicated, with the last cases being reported in 2006 and 1994, respectively. From 1978 to 2018, the total life expectancy for the 11 VPDs increased by 0.79 years.

**Conclusions:** China has had great success in controlling VPDs in recent decades, and the burden of major infectious diseases has been declining. The implementation of the National Immunization Program accelerated the decreases in the morbidity and mortality rates. Improving vaccination coverage is a key aspect of controlling diseases.

## Introduction

Vaccination has been one of the most successful and cost-effective public health interventions in the last century and has saved millions of people from various diseases(1). The benefits of vaccination include the eradication of one deadly disease, small pox, and the near eradication of another, poliomyelitis(2). In 1974, the World Health Organization (WHO) established the Expanded Program on Immunization (EPI) with the goal of increasing immunization coverage among children throughout the world(3). The EPI has contributed significantly to reducing the global burden of vaccine-preventable diseases (VPDs) over the past decades(4).

China has one of the largest and oldest immunization programs in the world, with over 16 million infants vaccinated each year(5). Work on immunization began in earnest as soon as the People’s Republic of China was founded in 1949(6). Based on immunization strategies, policy requirements and implementation progress, China’s immunization plan was divided into four stages from 1950 to 2018. The first stage was the pre-planned immunization program (1950-1977), during which the main strategy was to implement seasonal assault vaccinations of diphtheria vaccine (7), pertussis vaccine (8), Japanese encephalitis vaccine (9) and BCG vaccine (10). In 1978, in response to the call of WHO, the Ministry of Health proposed the concept of a Planned Immunization program suitable for China’s national conditions(11), thereby moving China’s vaccination work into the planned immunization stage (1978-2000)(12).

The immunization work made rapid progress during the immunization program stage (2001-2007). During this period, the cold chain system for immunization was established and improved, and the incidence of VPDs was controlled to a low level. In 2001, the Planned Immunization was re-organized and renamed the China National Immunization Program (NIP) to reflect work on a growing range of programmatic issues and the introduction of new vaccines(13). Increasing from the five vaccines used to prevent seven diseases during Planned Immunization, the NIP used 14 vaccines to prevent 15 diseases, including the routine vaccination of children with 11 vaccines to prevent 12 diseases(14).

Beginning in 2008, the state moved forward to the “Implementation Plan for Expanding the National Immunization Plan” (hereinafter referred to as “the Plan”), which signaled the beginning of the fourth stage (14). The overall goal of the Plan is to comprehensively implement an expanded NIP, continue to maintain a polio-free state, eliminate measles, control hepatitis B and further reduce the incidence of VPDs. According to the Plan, the vaccines against hepatitis B, BCG, polio, DPT, measles/DPT, hepatitis A, meningitis, encephalitis B and mumps are included in the NIP, and regular vaccination is carried out for school-age children.

In the past 70 years, China’s immunization program has been developed rapidly and comprehensively, making significant contributions in protecting population health, reducing the morbidity and mortality of VPDs and improving the per capita life expectancy. Recently, although some studies have reported on overall notification rates of VPDs, there has been no publication of systematic data on disease burdens associated with VPDs in China. Here, to quantify the impact of the national vaccination program on the prevention and control of infectious diseases, we evaluated patterns in morbidity and mortality of 11 VPDs in China based on national surveillance data. We also estimated the decrease of life expectancy caused by these infectious diseases since the introduction of their vaccines.

## Materials and methods

### Data sources

We constructed a standardized vaccine-preventable infectious disease dataset by extracting information from the National Disease Reporting System (NDRS). Data on the total population and natural deaths of the population were obtained from the China Statistical Yearbook. The compiled dataset included the number of reported cases and deaths and the computed incidence and mortality rates associated with the 11 VPDs for the period spanning 1950 to 2018. Data on morbidity and mortality stratified by demographic characteristics (e.g., age, sex, occupation) were not available at the time of the analysis.

Data on vaccination coverage of Japanese encephalitis, meningococcal meningitis and hepatitis A were collected from national vaccination coverage surveys that were conducted in selected provinces during 1998–2018. All surveys utilized multi-stage probability of selection proportional to population size sampling based on WHO guidelines(15). Data on vaccination rates for other diseases were obtained from the WHO website (http://apps.who.int/gho/data/node.main.A824?lang=en).

### Descriptive and statistical analyses

Eleven VPDs targeted by category 1 vaccines were selected: meningitis, measles, polio, diphtheria, pertussis, tuberculosis, hepatitis B, hepatitis A, Japanese encephalitis, mumps and rubella. The annual numbers of cases and deaths were divided by the respective population denominators to obtain annual incidence rates and mortality rates. Diseases were divided into two groups according to the transmission route (respiratory transmitted diseases and other diseases). To assess the historical incidence pattern of the 11 VPDs, their annual total incidence and mortality rates were calculated by standardizing the reported cases and deaths for each disease, with the total population of the country in that year taken as the denominator.

Information related to China’s NIP was obtained from various sources, including reports from the WHO Western Pacific Region Office (WPRO) and other published papers(16-19). We used the year during which each vaccine was introduced into the EPI as a cutoff year to define the pre-vaccination period (10 years prior to and including the cutoff year) and post-vaccination period (10 years after the cutoff year). The total number of years included in the analysis for each disease ranged from 8 to 20 years, depending on the data availability and cutoff year. By assuming that any changes to the incidence rates of the examined VPDs were entirely attributable to immunization, we estimated the effectiveness of the EPI by using quasi-Poisson regression models to estimate the change in the incidence rate for each disease from the pre-to post-vaccination period. Diseases without available data prior to integration of the vaccine in the EPI (i.e., the pre-vaccination period) could not be included in these analyses. The abbreviated life table method was used to calculate the decrease of life expectancy caused by infectious diseases from 1978 to 2018, and the distribution of these reductions in different age groups in 2010 and 2016. We were unable to obtain the incidence data for each age group from 1978 to 2003, and thus used the incidence ratio of each age group in 2004 for our calculation. The incidence ratio of each age group in 2017-2018 was calculated according to the incidence ratio in 2016. The potential years of life lost (PYLL) and potential working years lost caused by premature death from infectious diseases from 2004 to 2016 were estimated. Premature death was defined as death caused by not living to the expected lifespan, and was calculated based on the natural mortality rate of the Chinese census.

### Statistical software

We used R3.5.1 to construct quasi-Poisson regression models, and we used ArcGIS version 10.2 (ESRI, Redlands, CA, USA) to draw the thematic map of the disease incidence in different provinces.

## Results

### Overall patterns of the 11 VPDs

The greatest number of VPDs since 1950 was reported in 1959, when there were more than 11.5 million reported cases and a total incidence rate of 1723 per 100,000 persons (Figure 1); about 95.25% of these cases were made up of measles and pertussis, accounting for the large number of infected measles patients in China in 1960s. As immunization gradually progressed, the incidence rate of VPDs declined throughout the 1980s until it reached a nadir of 13 per 100,000 persons in 1989. The establishment of the National Notifiable Diseases Network Direct Reporting System (NNDNDRS) in 2004 led to the discovery of more cases, likely explaining why the overall incidence of the 11 VPDs has been on the rise since then. In general, the annual incidence of the 11 diseases showed a significant downward trend during the first three periods, especially during the EPI and NIP periods (Figure 1). In the fourth period, the incidence of the 11 diseases was basically stable at a relatively low level. In addition to our analysis of the 11 VPDs, we separately examined the long-term trends in a subset of six VPDs that had been continuously monitored throughout the study period (Japanese encephalitis, diphtheria, meningococcal meningitis, measles, poliomyelitis and pertussis; herein called the “long-studied” diseases or VPDs). The incidence of the six long-studied diseases was the same as that of the 11 diseases before 1990, because the data for the other five diseases were collected after 1990. Unlike the incidence of the 11 diseases, that of the six long-studied diseases was low in 1990 and has remained relatively stable since then.

**Figure 1.**
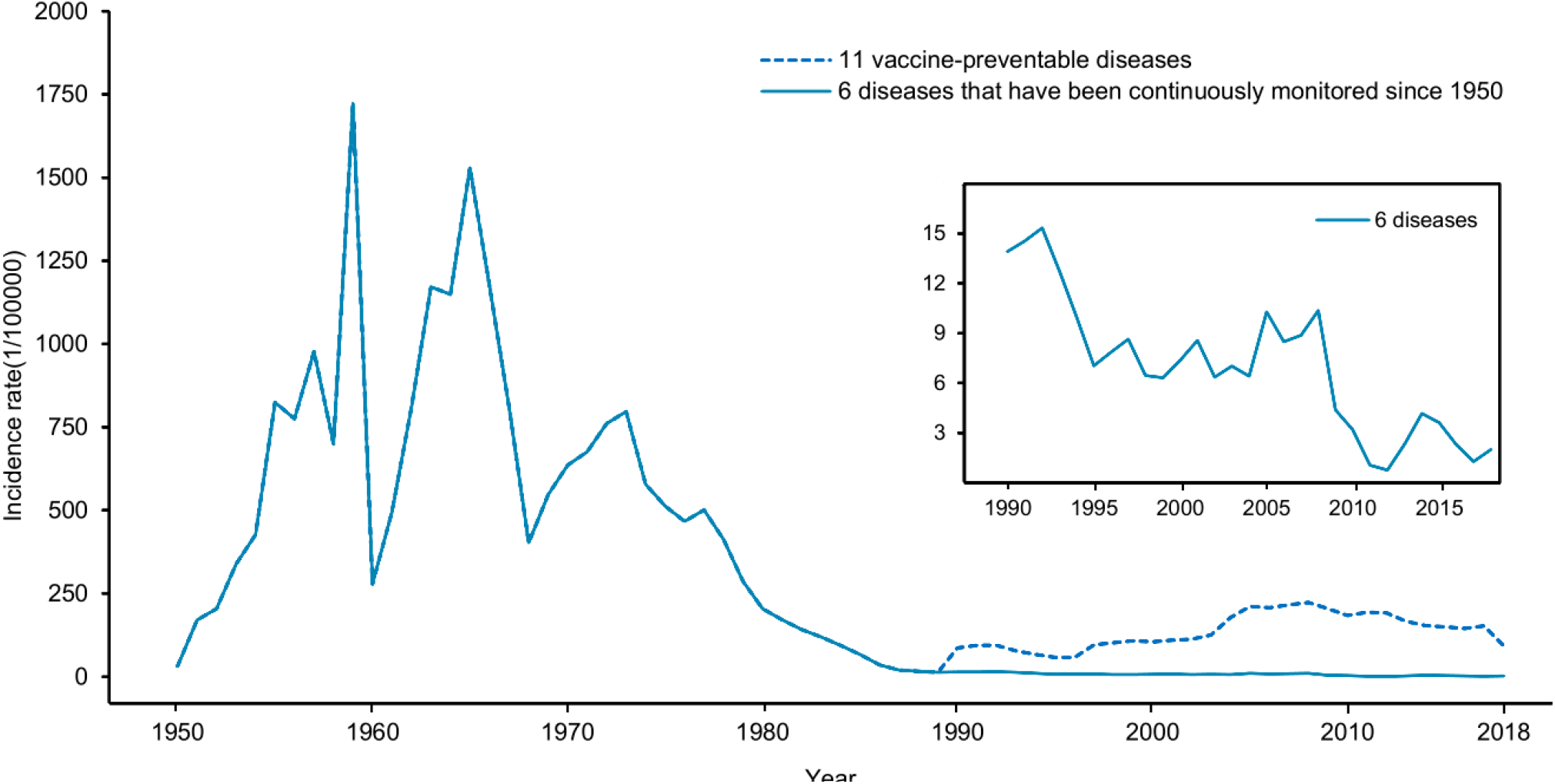
Annual incidence rate of the 11 selected vaccine-preventable diseases (VPDs) and six long-studied in China from 1950 to 2018. The dotted line in the large picture shows the incidence rate of the 11 VPDs and the solid line represents the six diseases that have been continually monitored since 1950 in China (herein called long-studied diseases: Japanese encephalitis, diphtheria, meningococcal meningitis, measles, poliomyelitis, pertussis). Since the incidence rates of the remaining five diseases were only available after 1990, the two lines overlap before then. The chart shows the incidence of the six long-studied diseases from 1990 to 2015. The small picture shows the incidence rate of these six diseases during 1990-2015.

Similar to the incidence data, the highest annual mortality rate of the 11 diseases was observed in 1959 (during the pre-planned immunization period), when the number of total deaths exceeded 335,700 cases. The highest annual mortality rate was 49.96/100,000 in 1959; the lowest annual mortality rate was 0.038/100,000 in 2017, and represented a decline of 99.92% compared with the 1959 rate. The annual mortality rate of the 11 diseases showed significant declines in different periods, with the most significant declines (nearly 100%) seen during the periods associated with the relevant immunization programs (Figure 2). Similarly, our separate analysis of the mortality trends for the six long-studied diseases revealed that, prior to 1990, the mortality rates for these six diseases were the same as those for the 11 diseases. After 1990, the mortality rates for the six diseases declined slowly and eventually approached zero.

**Figure 2.**
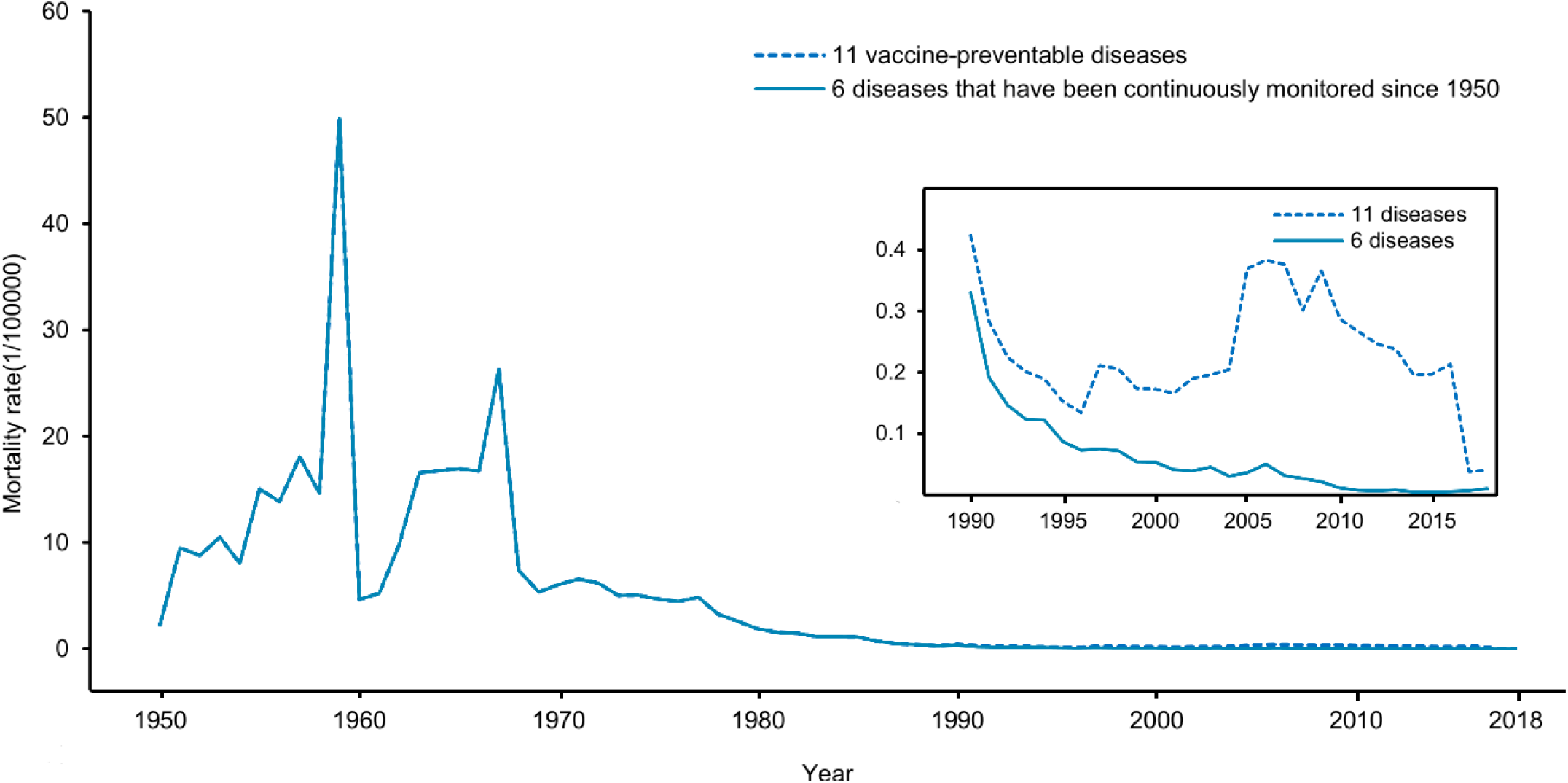
Annual mortality rate of 11 selected VPDs and six long-studied diseases in China from 1950 to 2018. The dotted line represents the mortality rate of the 11 VPDs and the solid line represents the six long-studied diseases. Since the mortality rates of the remaining five diseases were only available after 1990, the two lines overlap before that year.

Next, the 11 VPDs were classified into two groups according to their transmission routes. The results (Figure 3) showed that incidences of most of the 11 VPDs (respiratory-transmitted diseases and other diseases) peaked before 2000, whereas the highest notification rates were observed in the most recent year for hepatitis B and tuberculosis. The latter finding may reflect that the diagnostic level and detection rates of these diseases are currently at an all-time high. Moreover, the incidence of hepatitis B and tuberculosis has always been high in many areas, and there are many latent-infected people.

**Figure 3.**
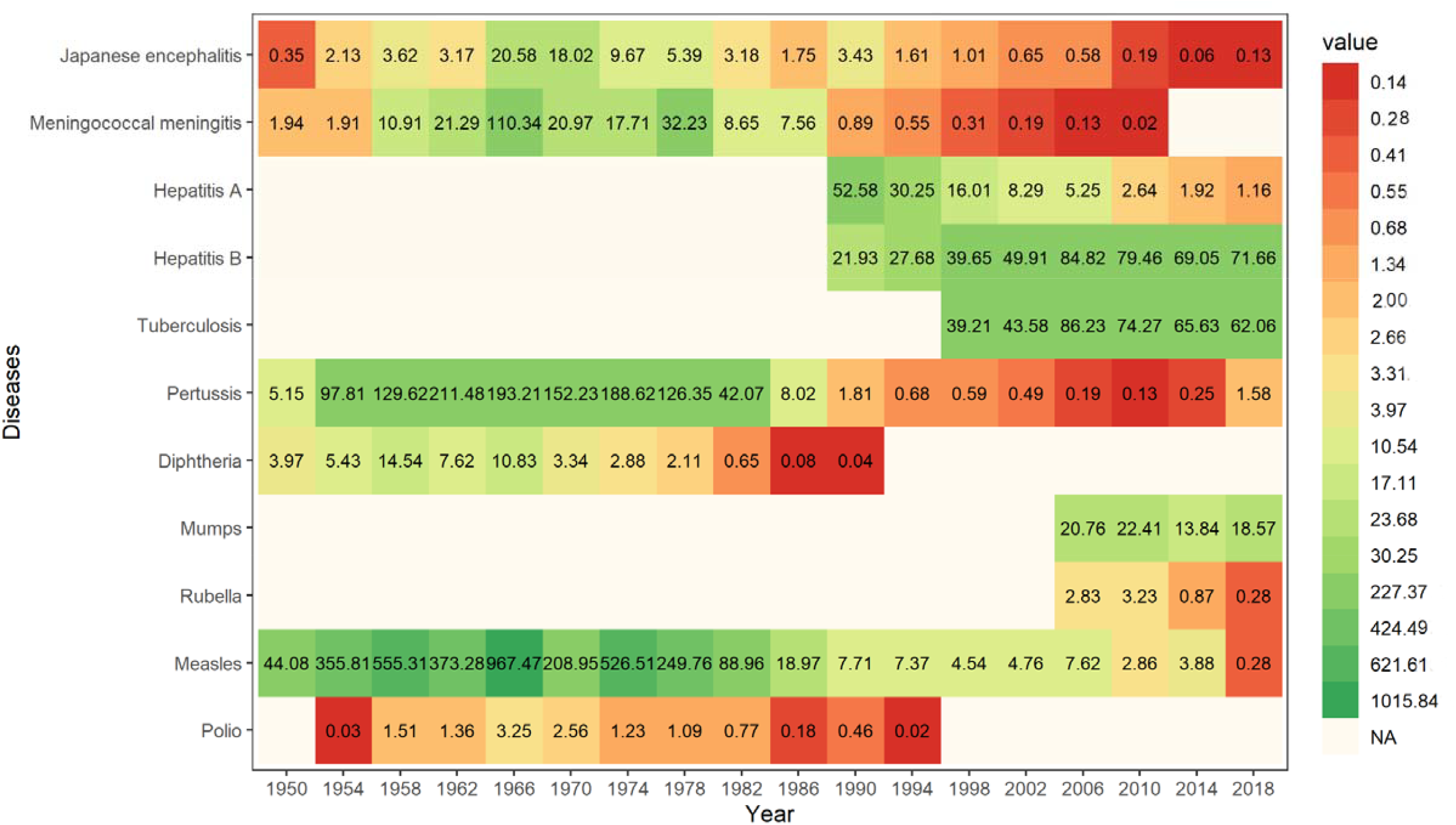
Annual incidence rates of 11 VPDs in China, 1950-2018.

### Types of diseases that have caused major health hazards at different stages

Based on the different stages of the immunization program in China, we selected 1977, 2001, 2008 and 2018 as the time nodes for the different periods, and compared the numbers of cases and deaths for the 11 diseases on these years. Figure 4 shows the diseases that ranked top five in the numbers of cases and deaths for each of the selected years. In 1977, out of the 11 studied diseases, the three highest numbers of cases were seen for measles (55.42%), pertussis (30.53%) and meningitis (11.85%), which was consistent with the highest incidence of measles and pertussis (95.25%) discussed in the first part of the Results section. During this period, the top three diseases that caused the most deaths were meningitis (51.05%), measles (25.35%) and Japanese encephalitis (15.24%). These findings mainly reflected the high incidences of these three diseases.

**Figure 4.**
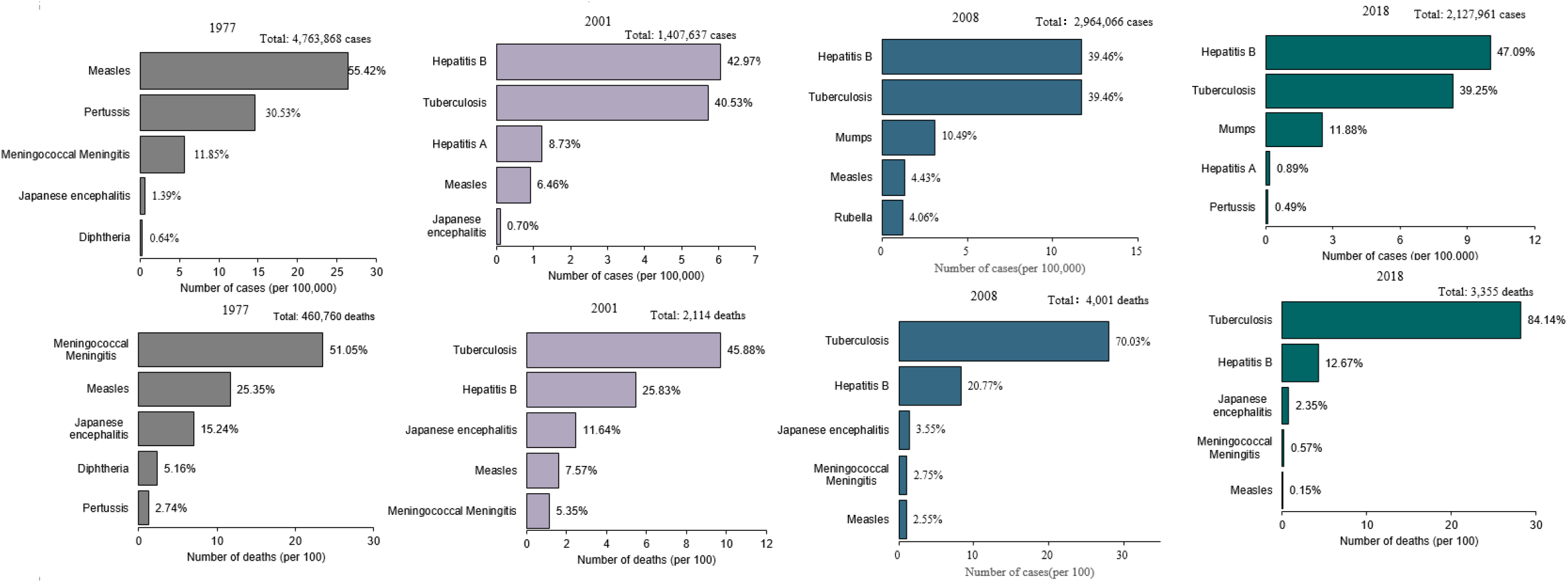
Top five VPDs in annual incidence and death rate in 1977, 2001, 2008 and 2018. The figure shows the incidence and death order of the top five diseases by year. The number of cases is the number of cases per 100,000 people, and the number of deaths is the number of deaths per 100 people. The percentiles in the figure represent the proportion of cases that occurred or died relative to the 11 diseases, and the numbers in the upper right corner of the figure indicate the total number of diseases or deaths in that year. No data were available for hepatitis B or TB in 1977.

In 2001, of the 11 studied diseases, the three highest numbers of cases came from hepatitis B (42.97%), tuberculosis (40.53%) and hepatitis A (8.73%), while the three highest numbers of deaths came from tuberculosis (45.88%), hepatitis B (25.83%) and Japanese encephalitis (11.64%). During the relevant period, China included the group A meningitis vaccine and the Japanese encephalitis vaccine into the planned immunization vaccination program. With the realization of the targeted 85% vaccine coverage for children in provinces, counties and townships in 1990, the types of diseases causing major health hazards have also changed. During the immunization program period, the hepatitis B (in 2002) and hepatitis A (in 2008) vaccines were included in the NIP. Thus, it is unsurprising that in the incidence sequence map of the 11 VPDs in 2018, hepatitis A had fallen from the top three and its case ratio had decreased significantly to 0.89%. During the period of the expanded immunization program, the Chinese government carried out a series of immunization-strengthening activities; these included nationwide measles immunization-strengthening activities, which were put into place in 2010 and made significant progress toward eliminating measles. The incidence of measles moved from fourth place (accounting for 4.43%) in 2008 to 0.18% in 2018.

### Total incidence of the six long-studied diseases by province

We collected data on the incidence of the six long-studied diseases in various provincial administrative districts from 1950 to 2016 and calculated the total incidence for each disease in each province for each of the four stages. As shown in Figure 5, the incidence of all six diseases was highest during the first stage, when China’s newly begun immunization planning was mainly based on the implementation of assault vaccination. Thereafter, the immunization program gradually took shape, as did the child immunization-planning framework. Since 1978, children’s program immunization work has been carried out nationwide, with timely and effective vaccinations administered to school-age children in accordance with the prescribed immunization procedures. The vaccines against BCG, polio, DTP and measles were included in the early immunization program for children in China. It is therefore unsurprising that, compared with the first stage, the incidence of all six diseases was much lower during the second stage. Interestingly, the incidence of these six diseases remained relatively high in many midwestern provinces compared to the eastern provinces. In the third and fourth stages, the by-province morbidity of the six long-studied diseases continued to decline; this was particularly notable in some provinces in the eastern and central regions, where the overall incidence of these diseases had dropped to less than 5/100,000 by 2016.

**Figure 5.**
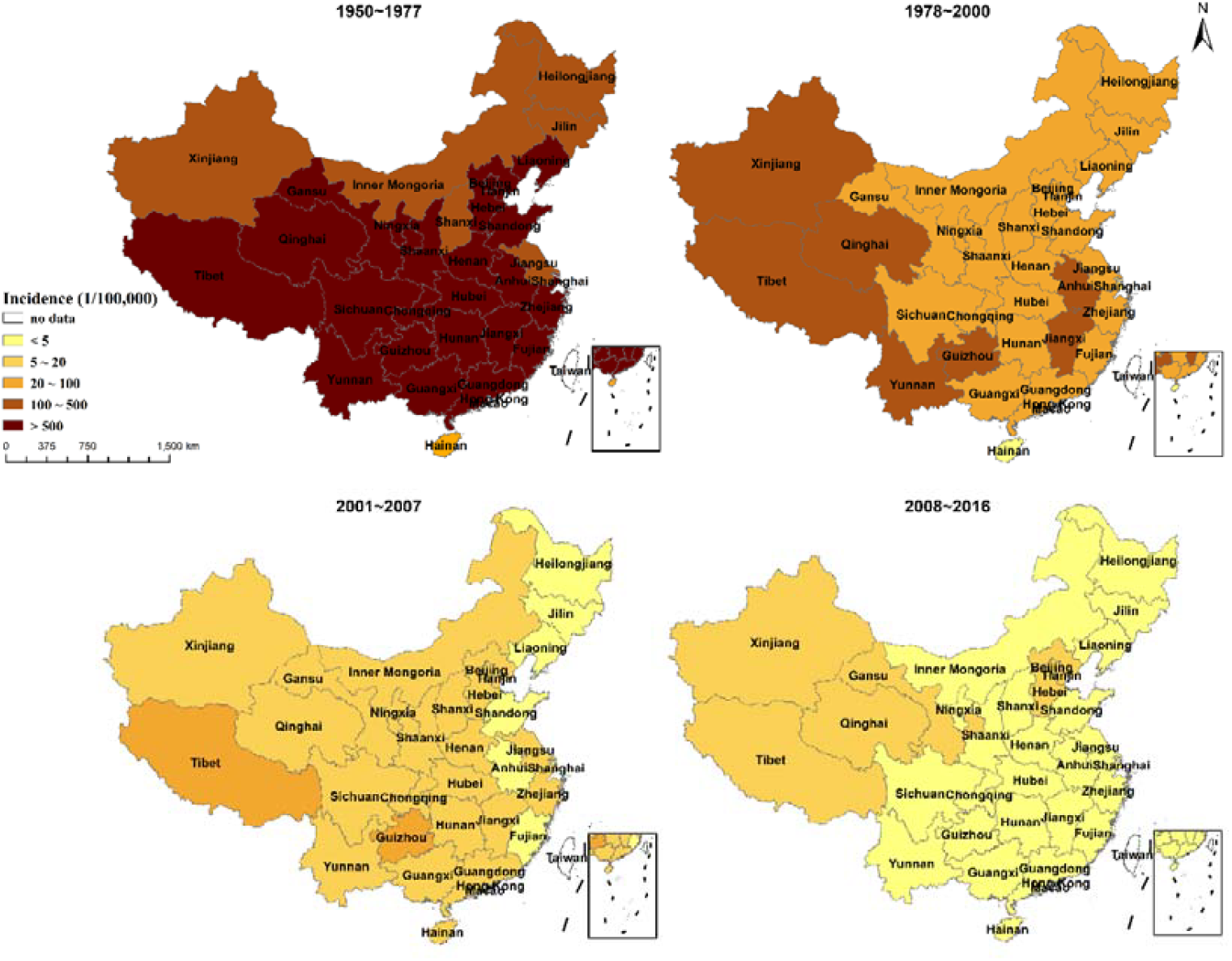
The total incidence of six long-studied diseases in different provinces in China since 1950. National distribution of the incidence rates of the six long-studied diseases; the darker the color, the higher the incidence rate. Chongqing became a municipality directly under the central government in 1997; prior to that, it belonged to Sichuan Province, and thus disease epidemic data were not listed separately until 1997. When calculating the incidence before 1997, we used the incidence rate of Sichuan Province. In addition, sub-regional information for the 11 diseases was only collected in 2016.

### Impact of vaccination coverage on morbidity rates of VPDs

Figure 6 shows the relationship between the vaccination coverages and incidence rates of the VPDs. Focusing on diphtheria and pertussis, we see that after the combined DTP vaccine became available free for all children across the country in 1978, the transmission of the two diseases declined significantly. Similarly, the incidence of measles rapidly declined shortly after the measles-rubella vaccine and the measles-mumps-rubella vaccine were integrated into the EPI program in 1978 and 2007, respectively, and also following the initiation of supplementary immunization activity in 2008 (Figure 6). Conversely, even though effective vaccines against tuberculosis and hepatitis B were integrated into the EPI in the 2000s, the incidences of these diseases have continued to rise. The incidence trends of the other nine diseases were basically consistent with those of the corresponding vaccination coverages. In 1982, the Ministry of Health set EPI targets (to be reached by 1990) of 80%-90% coverage for DPT and 90%-95% coverage for OPV and measles by the time a child reached school age. Therefore, the coverage of these vaccines increased during the 1980s. By 2003, however, central government funds accounted for only 1% of total immunization expenditures and more than 50% of expenditures were being made at the village and township levels, with the result that the coverage of vaccines was on the decline. In 2004, the central government for the first time allocated funds for vaccine delivery and administration at the village and township levels. In 2004–2005, the State Council passed the Law on the Prevention and Treatment of Infectious Diseases, reaffirming the requirements for all children to be fully vaccinated. Together, these measures strengthened the financing for vaccines and vaccination services, resulting in increased and more equitable coverage and a historically low VPD burden. From 1978 to 2018, the incidence of pertussis fell 98% from 126.35 to 1.58 per million; measles fell 99% from 249.76 to 0.28 per 100,000; meningococcal meningitis fell 99% from 32.23 to 0.01 per million; and JE fell 98% from 5.39 to 0.13 per million. The last case of diphtheria was reported in 2006.

**Figure 6.**
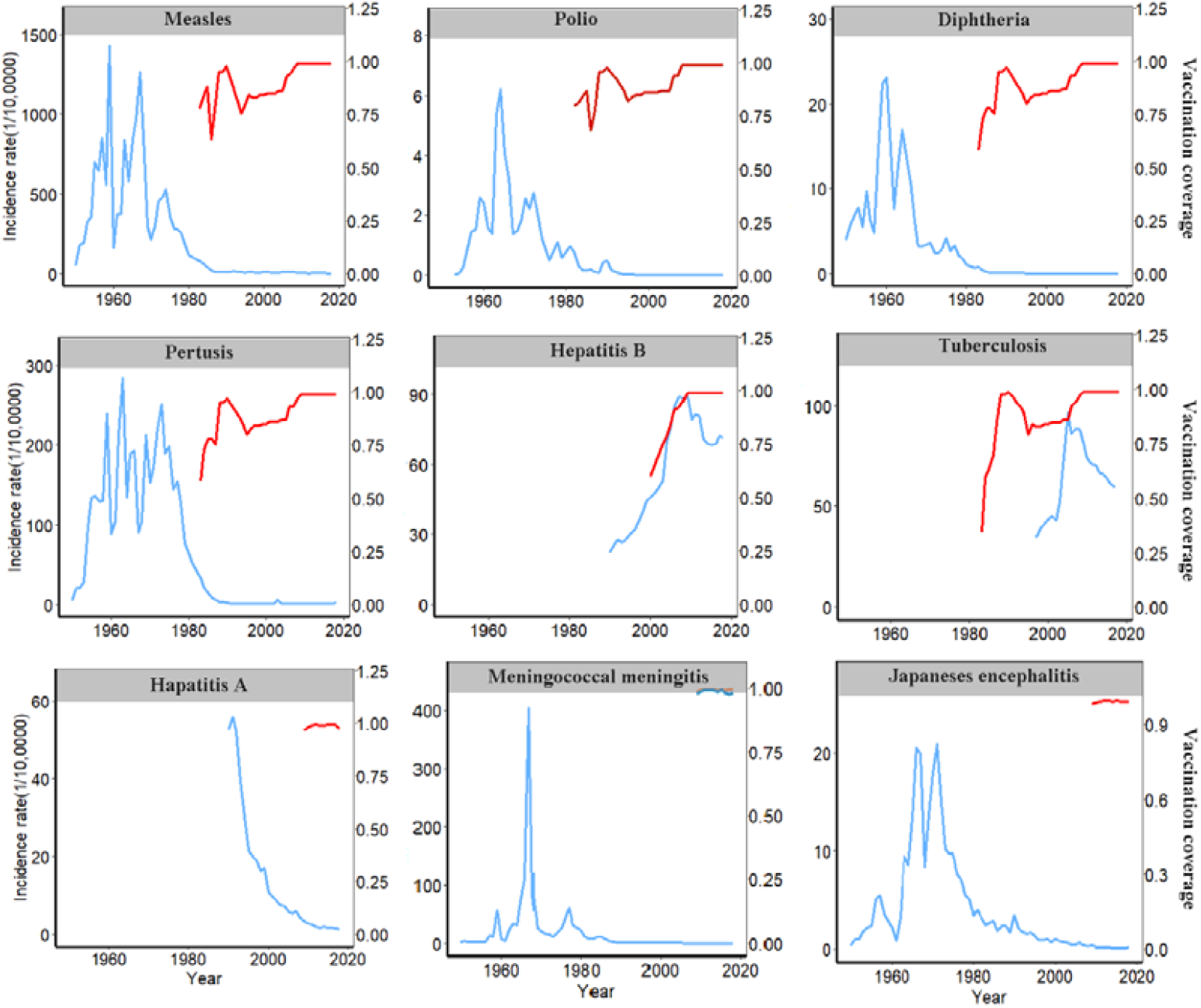
Annual incidence rates of selected VPDs and vaccination coverage. The gray background indicates the period in which the vaccination for a given disease was integrated into the national Expanded Program on Immunizations (EPI). The blue line indicates the morbidity rate of the disease; the red line represents the vaccination rate of the disease; and the green line represents the vaccination rate for the group A + C vaccine. Data on vaccination coverage of Japanese encephalitis, meningococcal meningitis and hepatitis A were collected from national vaccination coverage surveys. Data on vaccination rates for other diseases were obtained from the WHO website (http://apps.who.int/gho/data/node.main.A824?lang=en).

### Quasi-Poisson regression model

As shown in Table 1, the results of the quasi-Poisson regression model revealed that, with the exceptions of tuberculosis (for which we lacked epidemic data before 1997), hepatitis B (the incidence increased), mumps and rubella (the confidence interval included zero), the incidence of all diseases decreased significantly after the relevant vaccine was introduced. For example, the incidence of measles decreased by 79.66% in the 20 years before and after the introduction of the vaccine, and whooping cough, diphtheria and polio decreased by 82.60%, 81.39% and 74.50%, respectively. The inclusion of vaccines in immunization programs was also associated with changes in disease mortality, with measles mortality falling by 81.62%, whooping cough by 81.42%, diphtheria by 80.08% and polio by 67.42%. Hepatitis B exhibited significant increases in morbidity and mortality when we compared the numbers before and after introduction of the vaccine; however, we speculate that this may be due to underreporting during the earlier stages.

**Table 1.** Impact of routine vaccination on the incidence rates of the selected vaccine-preventable diseases.

| **Disease** | **Year of integration into the EPI** | **Time period\* (years)** | **Changes in incidence (%) (95% confidence interval)** |
| --- | --- | --- | --- |
| Measles | 1978 | 20 years<br>(1969-1988) | -81.63<br>(-69.94, -88.78) |
| Pertussis | 1978 | 20 years<br>(1969-1988) | -83.01<br>(-73.03, -89.29) |
| Diphtheria | 1978 | 20 years<br>(1969-1988) | -81.95<br>(-68.94, -89.51) |
| Polio | 1978 | 20 years<br>(1969-1988) | -74.52<br>(-52.11, -86.44) |
| Tuberculosis<sup>†</sup> | 1978 | N/A | N/A |
| Hepatitis B | 2002 | 20 years<br>(1993-2012) | 109.29<br>(77.92, 146.20) |
| Mumps<sup>‡</sup> | 2007 | 8 years<br>(2004-2011) | 28.17<br>(0.62, 64.25) |
| Rubella<sup>‡</sup> | 2007 | 8 years<br>(2004-2011) | 81.58<br>(-11.50, 272.57) |
| Hepatitis A | 2007 | 10 years<br>(2003-2012) | -54.85<br>(-37.27, -63.18) |
| Meningococcal meningitis | 2007 | 10 years<br>(2003-2012) | -78.55<br>(-51.29, -90.04) |
| Japanese encephalitis | 2007 | 10 years<br>(2003-2012) | -58.87<br>(-38.11, -72.67) |

\*The number of years included in the regression is different for each disease and is based on the data availability and the year of integration into the EPI.
<sup>†</sup> Tuberculosis: data are available from 1997 onward.
<sup>‡</sup> Mumps and rubella: data available from 2004 onward.

### The burden of the 11 VPDs

From 1978 to 2018, the total increase in the life expectancy at death due to the 11 VPDs was 0.791 years (about 9.25 months) (Figure 7). The increases by disease included: meningitis, 2.86 months; JE, 1.95 months; measles, 1.83 months; hepatitis A, 0.09 months; and hepatitis B, 0.50 months. For all 11 VPDs, the value of this indicator was 0.01 months in 2018 and 1.30 months in 1978. By 2018, the vaccines had increased the life expectancy for these 11 diseases by 1.29 months compared to the data from 1978 As shown in Figure 7, since the implementation of the planned immunization program, the increased life expectancy without the cause of death of 11 VPDs has decreased rapidly in years. Compared to the 1978 value of 0.11, the increased life expectancy without the cause of death for the 11 VPDs had decreased by 99.1% in 2018.

**Figure 7.**
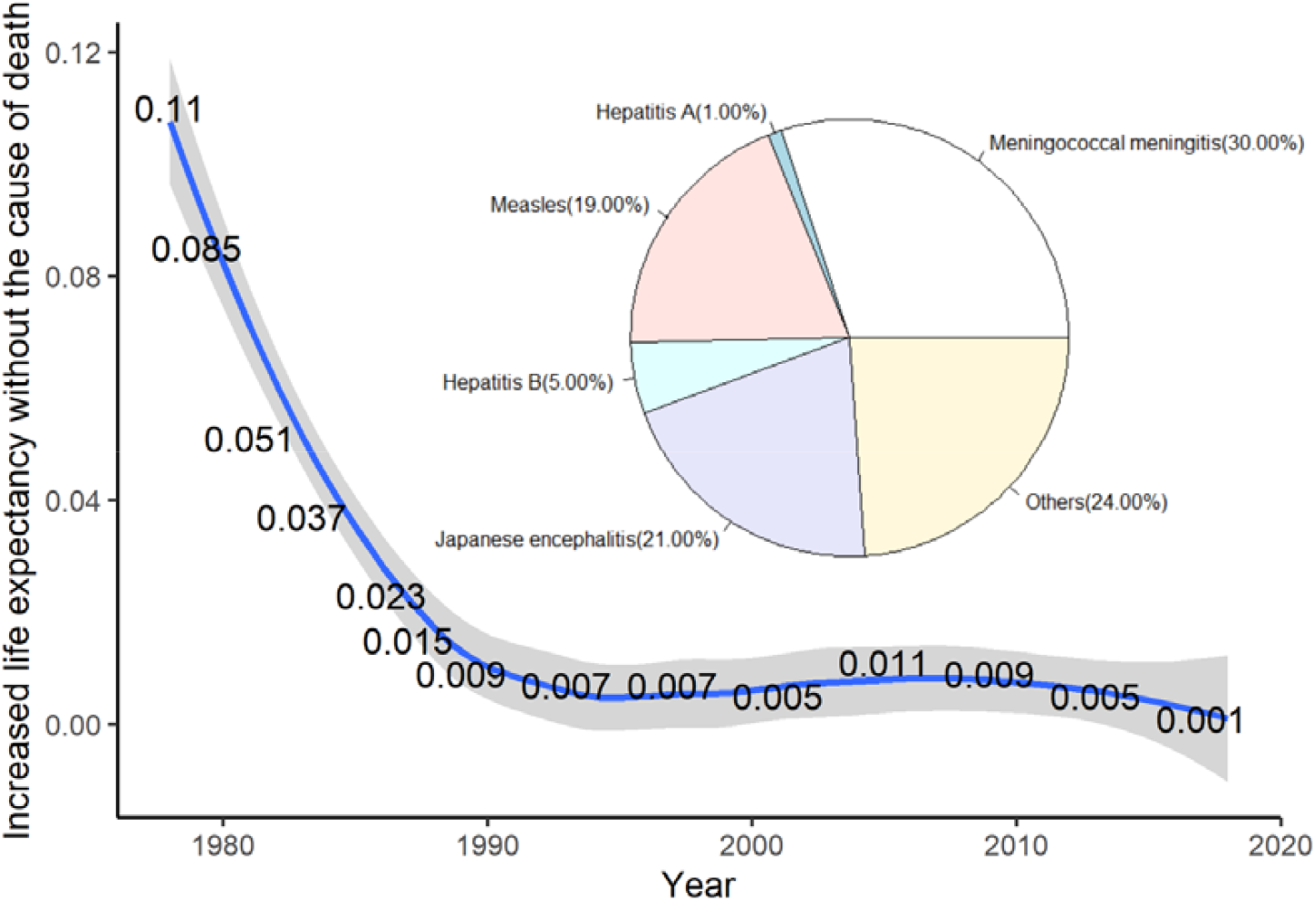
The decrease of life expectancy caused by infectious diseases from 1978 to 2018. The figure shows the impact of the 11 studied VPDs on life expectancy from 1978 to 2018(The lines were smoothed by using locally estimated scatterplot smoothing (LOESS) regression). The life expectancy from 2004 to 2018 was calculated from the census data of China in 2010. Since we did not have data for the third and fourth censuses, which were conducted in 1982 and 1990, the life expectancy from 1978 to 2003 was calculated according to the census data of China in 2000. We were unable to obtain the incidence data of each age group for measles, meningococcal meningitis and Japanese encephalitis from 1978 to 2003 and for hepatitis A and hepatitis B from 1990 to 2003. We used the incidence ratios of each age group in 2004 for our calculation. The incidence ratio of each age group in 2017-2018 was calculated according to the incidence ratio in 2016. For better comparison, the age-specific proportion of each disease was calculated as the proportion of the 11 studied diseases.

The impacts of these infectious diseases on life expectancy were also examined for different age groups. Our results revealed that the impacts of the 11 VPDs gradually decreased with time in people over 15 years of age (Suppl. Fig. S1 and Suppl. Fig. S2), as well as in younger age groups.

For hepatitis A and hepatitis B, which were not included in the EPI in 1978, the impact on life expectancy has been calculated since 1990, when these vaccines were introduced. When the hepatitis B vaccine was first introduced in 1990, the impact of hepatitis B-related death on the life expectancy of newborns was 0.023 months; in 2018, when the hepatitis B vaccination rate reached more than 95%, the impact of hepatitis B-related death on the life expectancy of newborns dropped to 0.007 months. Compared with the value in 1990, the contribution of the hepatitis B vaccine to the life expectancy of newborns in 2018 was 0.16 months. Similarly, the 1990 introduction of the hepatitis A vaccine increased the life expectancy of newborns in 2018 by 0.015 months. By 2018, the life expectancy of neonates for meningococcal and Japanese encephalitis increased by 0.53 months and 0.23 months, respectively, relative to the 1978 levels.

When the measles vaccine was first added to the EPI in 1978, the impact of measles death on the life expectancy of newborns was 0.40 months. In 2018, when the measles vaccine vaccination rate was above 95%, the death rate impact was 0.00002 months. The impact of several other VPDS on life expectancy in 1978-2018 is presented in the supplementary information. Overall, the impact of these 11 diseases on life expectancy decreased during the study period.

Similar results were obtained for the loss of healthy life years and loss of work years due to early deaths from these infectious diseases. Figures 10 and 11 show the loss of healthy life years and loss of work years due to 11 VPDs. From 2004 to 2016, the total loss of healthy life years due to early death caused by the 11 VPDs was 1,182,322 person-years. As shown in Figure 10, except for an increase seen in 2012, the loss of healthy life showed a downward trend over time. In 2016, the loss of healthy life years due to early death from the infectious diseases was 53,136.77 person-years, which was significantly lower than the 81,592.79 person-years in 2004 (a decrease of 37.5%).

## Discussion

During the study period, the greatest number of VPDs was reported in 1959; in this year there were more than 11.5 million cases and a total incidence rate of 1723 per 100,000 persons (Figure 1). Of these cases, about 95.25% comprised measles and pertussis; this was subsequently reflected in the large number of infected measles patients seen in China in the 1960s. These high rates mainly reflect the lack of any large-scale vaccination against measles and pertussis in China at that time.

Disease control has always been the main goal of China’s immunization efforts, and disease incidence has always been the main metric used to guide the development of immunization strategies and assess immunization program performance(6). Here, we explored patterns in the morbidity and mortality of 11 VPDs in China based on national surveillance data, with the goal of evaluating the effect of China’s immunization programs on the prevention and control of infectious diseases. The total incidence rate of the 11 VPDs dropped significantly from 1950 to 2018, with a particularly notable decrease seen after the 1970s. However, the changes of the total incidence for the 11 VPDs differed among the four stages of China’s immunization program. The total incidence increased between 1950 and 1965 but has largely declined since 1966. After 1990, the total incidence increased slightly, perhaps in response to the establishment and improvement of the National Notifiable Diseases Network Direct Reporting System. We also selected six of the 11 diseases for further analysis, as they had been continuously monitored since 1950. The total incidence trend for the six long-studied diseases was similar to that of the 11 studied diseases, except that the incidence of the former had been stable at a low level since 1990. Compared with this difference in the morbidity trends of the 11 studied diseases versus the subset of the six long-studied diseases, both groups showed similar changes of mortality, which rose from 1950 to 1965 and then declined after 1966. The top five VPDs in terms of annual incidence and death rate were identified, and this analysis showed that the proportions of deaths caused by meningococcal meningitis, Japanese encephalitis and pertussis had substantially declined by 2018.

Successful implementation of routine immunization programs against VPDs has contributed to a steep decrease in the incidence of these diseases in China. The national smallpox vaccination campaign in 1949-1952 used more than 500 million doses of vaccine and led to a vaccination coverage of >90% nationwide and the elimination of the disease in China in the 1960s (20-22). Routine vaccination for prevention of polio, diphtheria, pertussis, tetanus, measles and tuberculosis was initiated in the 1960s in the larger cities and later expanded to rural areas, leading to substantial declines in the incidence and mortality of these diseases. The burden of hepatitis B in China remains much higher than that of many other infectious diseases, likely due to an increased awareness of the disease in the population; however this will soon be countered by the results obtained from hepatitis B vaccination, which was integrated into the national children’s routine vaccination program in 1992 (23, 24).

A study conducted in the US comparing trends of 13 VPDs showed declines greater than 92% in cases of diphtheria, mumps, pertussis and tetanus during 1980 to 2005 (25), which is completely in line with our present findings. In another study in the US, vaccination with seven recommended childhood vaccines was estimated to prevent 14 million cases of these diseases in every birth cohort(26). A US-based study on the long-term trends of weekly surveillance reports for eight VPDs estimated that about 103 million cases were prevented by vaccination(27). In other countries, various studies have highlighted the benefits of vaccination policies. Analysis of the incidence and mortality for five VPDs captured by Vietnam’s national surveillance suggested that between 1980 and 2010, up to 5.7 million disease cases and 26,000 deaths may have been prevented by the extended program of immunization(28). A study conducted in Iran demonstrated that vaccination has had a positive impact on the control of four communicable diseases: the death rates for diphtheria and tetanus decreased by nearly 85% from 1990 to 2010, while the fatality rate for measles declined by 94%(29).

A study by Janice C. Wright and Milton C. Weinstein (1998), published in *The New England Journal of Medicine*, found that neonatal hepatitis b vaccination contributed only 0.28 months to life expectancy, and that even the better effects of vaccines against measles, rubella, mumps and whooping cough did not exceed 0.11 months(30). Here, we report that, compared with the 1990 data, the contribution of hepatitis B vaccine to the life expectancy of newborns in 2018 was 0.16 months. Compared with the 1978 data, the contribution of the measles vaccine to the life span of newborns in 2018 was 0.39998 months. Similarly, compared with the period before the introduction of hepatitis A vaccine in 1990, the life expectancy of newborns increased by 0.015 months in 2018, while for those who joined EPI in 2007, compared with 1978, the life expectancy of newborns increased by 0.53 months and 0.23 months in 2018 (Suppl. Table S2). Thus, vaccination against the 11 VPDs studied herein had significant positive effects on life expectancy.

A few limitations of our study should be noted. First, the increasing incidence of notifiable diseases during the first few years following the implementation of national surveillance, particularly before 1960 (Figure 1), may not necessarily represent a rise in the occurrence of infections but rather a gradual improvement in the notification system (e.g., with more provinces reporting cases in the later years). Second, the passive nature of the surveillance system could have resulted in underreporting and/or under-ascertainment of the true disease burden. This is particularly true during periods when there was massive political, social and/or economic chaos nationwide, such as during the Cultural Revolution (1966-1976). Indeed, this could explain the sudden drop of incidence seen in the late 1960s (Figure 1). Aside from such societal changes, individual behavioral factors also need to be taken into consideration when we examine the underreporting of diseases. In the present study, we did not account for underreporting or uncertainty in the data analysis, and we acknowledge that this could have introduced bias, limiting our interpretation of patterns in the incidence of notifiable diseases in China.

## Data Availability

We constructed a standardized vaccine-preventable infectious disease dataset by extracting information from the National Disease Reporting System (NDRS). Data on the total population and natural deaths of the population were obtained from the China Statistical Yearbook. Data on vaccination coverage of Japanese encephalitis, meningococcal meningitis and hepatitis A were collected from national vaccination coverage surveys that were conducted in selected provinces. Data on vaccination rates for other diseases were obtained from the WHO website (http://apps.who.int/gho/data/node.main.A824?lang=en).

## Author contributions

J.P., Y.W. and W.W. designed and implemented the study. J.P., Y.W., Z.L. and Z.Y. collected the relevant data. J.P. and Y.W. analyzed and processed the results and drafted the manuscript. H.Y., J.Y., Y.W. Q.Z. and WW revised the structure of the paper and polished the language. All authors critically reviewed and approved the final version of the manuscript.

## Competing interests

The authors declare that they have no competing interest.

---

**Suppl. Fig. S1.**
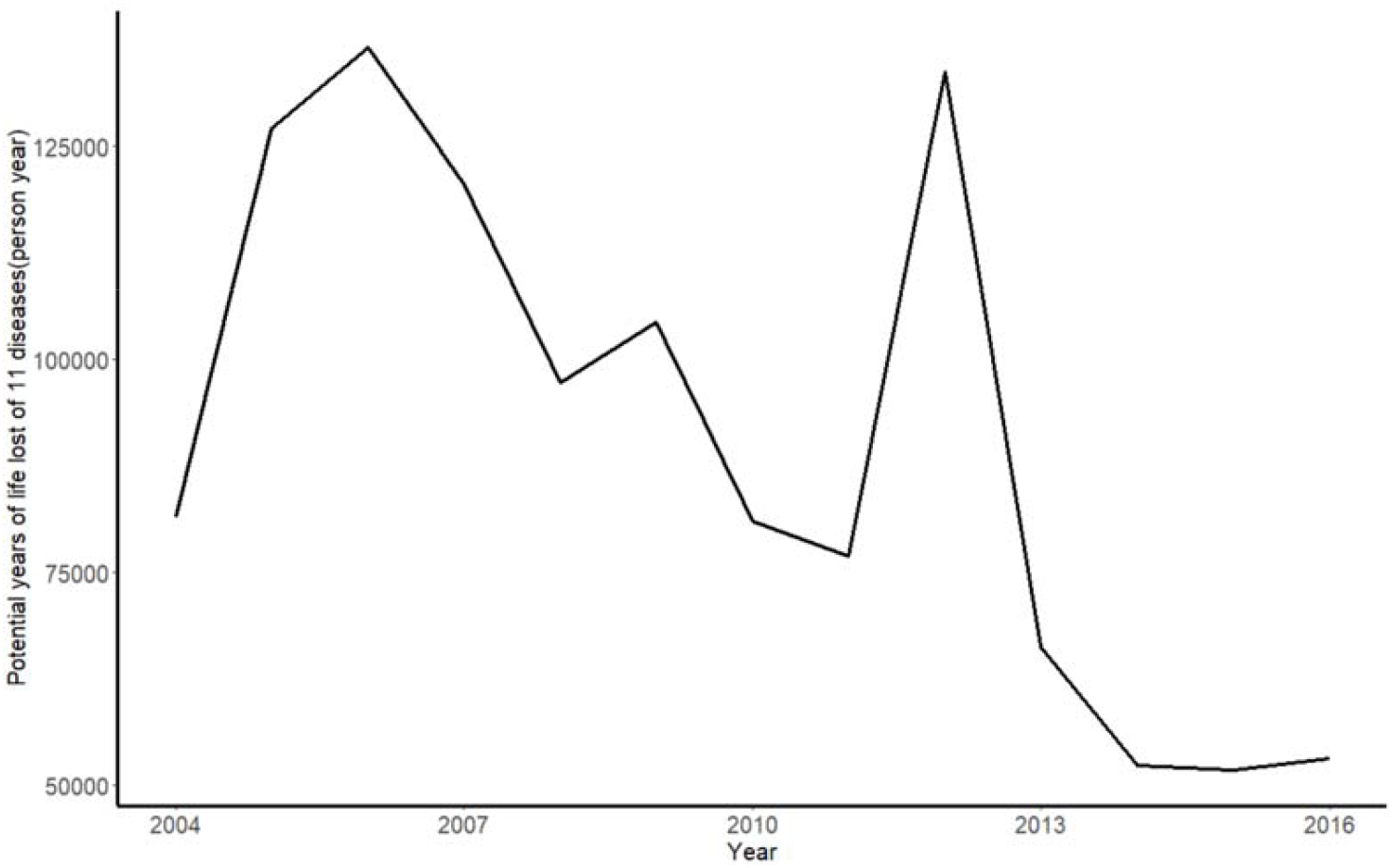
Potential years of life lost for the 11 diseases.

**Suppl. Table S1.** Loss of healthy life and work years due to early death from the 11 infectious diseases.

| Year | Loss of healthy life years<br>(person years) | Total<br>population | Early mortality<br>rate | Loss of work years<br>(person years) |
| --- | --- | --- | --- | --- |
| 2004 | 81592.79 | 1299880000 | 0.00006277 | 47410 |
| 2005 | 126949.6 | 1307560000 | 0.00009709 | 71137.5 |
| 2006 | 136593.4 | 1314480000 | 0.00010391 | 76960 |
| 2007 | 120585.1 | 1321290000 | 0.00009126 | 66375 |
| 2008 | 97181.57 | 1328020000 | 0.00007318 | 53487.5 |
| 2009 | 104323.7 | 1334500000 | 0.00007817 | 55740 |
| 2010 | 81033.93 | 1340910000 | 0.00006043 | 42905 |
| 2011 | 76899.41 | 1347350000 | 0.00005707 | 41262.5 |
| 2012 | 133739.4 | 1354040000 | 0.00009877 | 70355 |
| 2013 | 66203.94 | 1360720000 | 0.00004865 | 35052.5 |
| 2014 | 52293.87 | 1367820000 | 0.00003823 | 27477.5 |
| 2015 | 51788.21 | 1274620000 | 0.00004063 | 26907.5 |
| 2016 | 53136.77 | 1382710000 | 0.00003843 | 26955 |
| Total | 1182322 | 17333900000 | 0.00006821 | 642025 |

**Suppl. Fig. S2.**
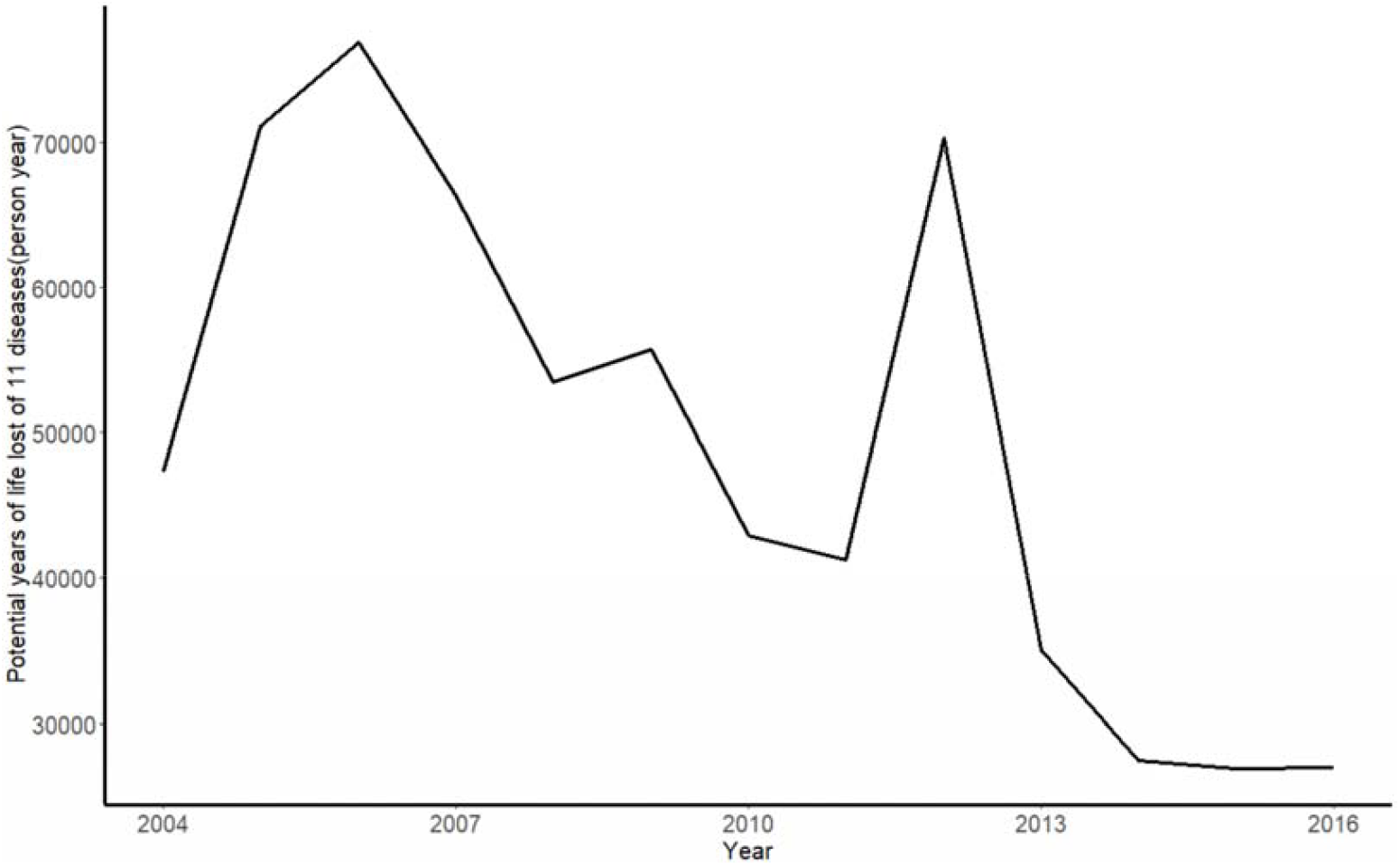
Disability-adjusted life years for the 11 diseases.

**Suppl. Table S2.**
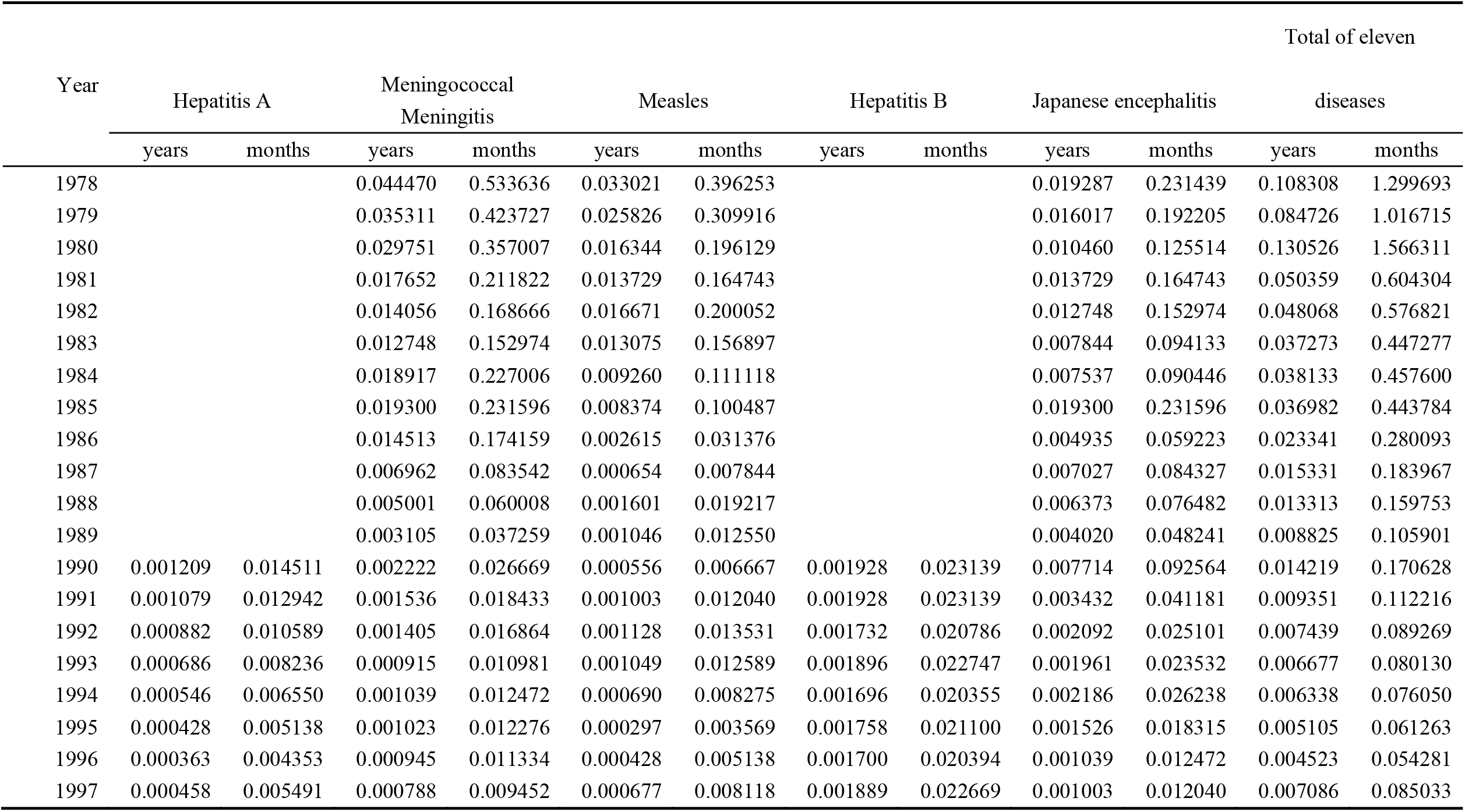
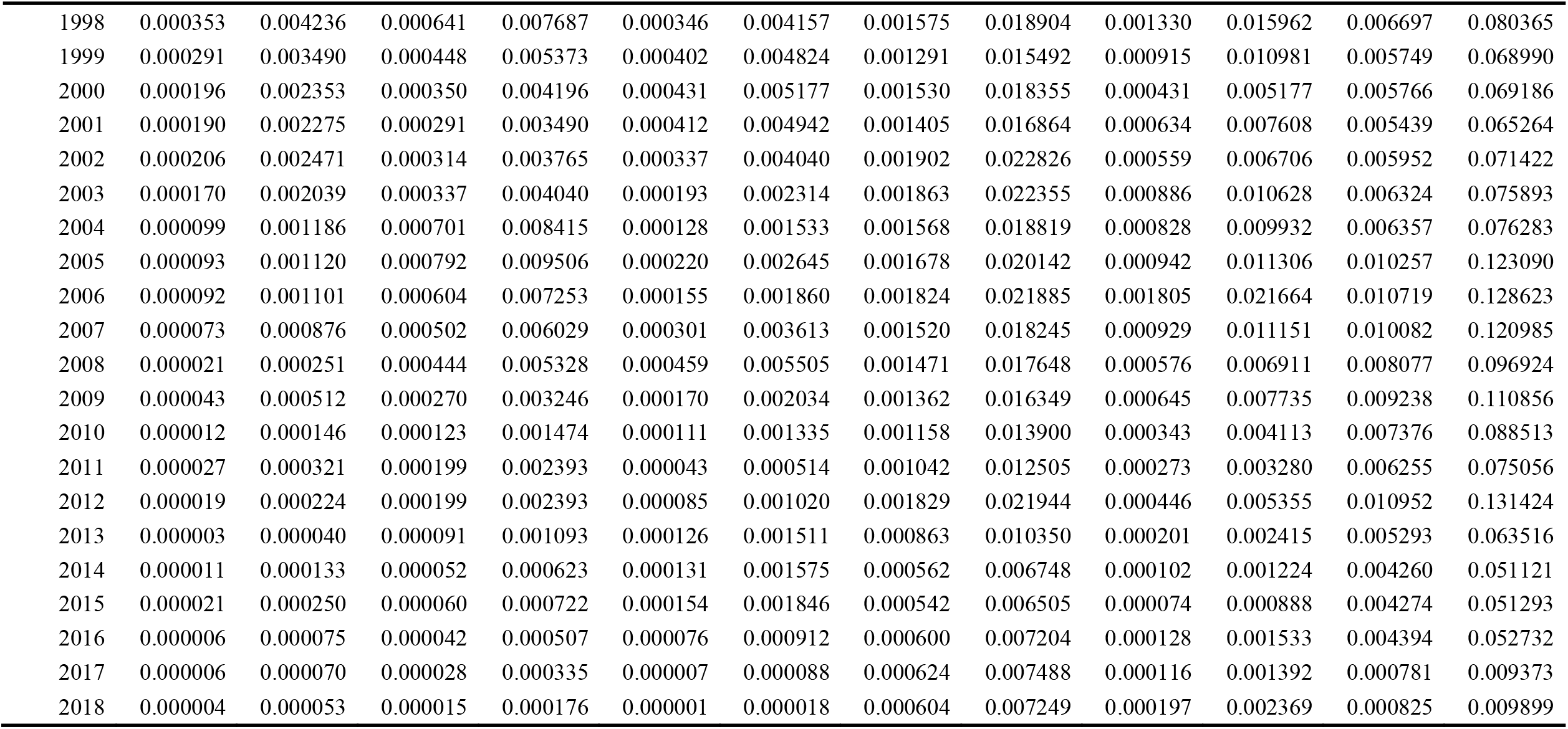
the effect of VPDS on life expectancy in 1978-2018.

## Notes

### Competing Interest Statement

The authors have declared no competing interest.

### Funding Statement

NONE

### Author Declarations

The Institutional Review Board at the School of Public Health, Fudan University, approved the study protocol with a waiver of informed consent. Data were analyzed at aggregate level and no participants were contacted.

